# Ablation of Apparent Diffusion Coefficient Hyperintensity Clusters and Mesial Temporal Lobe Epilepsy Improves Seizure Outcomes after Laser Interstitial Thermal Therapy

**DOI:** 10.1101/2022.04.01.22273326

**Authors:** Min Jae Kim, Brian Hwang, David Mampre, Serban Negoita, Yohannes Tsehay, Haris Sair, Joon Y. Kang, William S. Anderson

**Author notes:** Corresponding Author: Brian Y. Hwang, M.D., 18D, Department of Neurosurgery, Johns Hopkins School of Medicine, 600 N. Wolfe Street, Meyer 8-181, Baltimore, MD 21287.

## Abstract

**Objective:** MR-guided Laser Interstitial Thermal Therapy (LiTT) is a minimally invasive surgical procedure for intractable mesial temporal epilepsy (mTLE). LiTT is safe and effective but seizure outcomes are highly variable due to patient variability, suboptimal targeting, and incomplete ablation of epileptogenic zone. Apparent Diffusion Coefficient (ADC) is an MRI sequence that can identify potential epileptogenic foci in the mesial temporal lobe to improve ablation and seizure outcome. The objective of this study was to investigate whether ablation of tissue clusters with high ADC values in the mesial temporal structures is associated with seizure outcome in mTLE after LiTT.

**Methods:** Thirty mTLE patients who underwent LiTT at our institution were analyzed. Seizure outcome was categorized as complete seizure freedom (ILAE Class I) and residual seizures (ILAE Class II – VI). Volumes of hippocampus and amygdala were segmented from preoperative T1 MRI sequence. Spatially distinct hyperintensity clusters were identified in the preoperative ADC map. Percent cluster volume and number ablated were associated with seizure outcomes.

**Results:** The mean age at surgery was 36.6 years and mean follow-up duration was 1.9 years. Proportions of hippocampal cluster volume (35.20% vs. 16.5 %, p = 0.014) and cluster number (27.1 % vs 4.2 %, p = 0.0007) ablated were significantly higher in patients with seizure freedom. For amygdala clusters, only proportion of cluster number ablated was significantly associated with seizure outcome (13.2 % vs. 0 %, p = 0.016). Ablation of hippocampal clusters predicted seizure outcome, both by volume (AUC = 0.7679) and number (AUC = 0.8086) ablated.

**Significance:** Seizure outcome after LiTT in mTLE patients was significantly associated with the extent of cluster ablation in the hippocampus and amygdala. The results suggest that preoperative ADC analysis may help identify high-yield pathological tissue clusters that represent epileptogenic foci. ADC based cluster analysis can potentially assist ablation targeting and improve seizure outcome after LiTT in mTLE.

## Introduction

Mesial temporal lobe epilepsy (mTLE) is one of the most prevalent types of seizures in the world. Up to a third of the mTLE patients become drug resistant and require surgical intervention to control their seizures. MRI-guided Laser Interstitial Thermal Therapy (LiTT) is a minimally invasive surgical procedure that is rapidly becoming an alternate first-line surgical option in drug-resistant mTLE.^2^ LiTT allows for more selective targeting and has been associated with shorter hospital stay^2, 3^ and reduced postoperative neuropsychological complications compared to anterior temporal lobectomy, which remains the gold standard.^4, 5^

While LiTT is promising, post-operative seizure outcome is highly variable among patients and centers. The inconsistent outcome may be partly due to variation in mesial temporal love anatomy as well as in the size and location of the epileptogenic foci that current practice does not account for. Past studies have proposed several predictors of seizure outcome after LiTT, including target structures,^6, 7^ ablation volumes,^7^ and surgical approaches.^6, 8, 9^ However, these factors remain controversial in the literature with conflicting results across centers. Since LiTT is designed to target a smaller, localized tissue with real-time MR guidance, it is imperative to identify pre-operative variables that can help surgeons identify precise location of target tissues that will lead to meaningful improvement in long-term seizure outcome.

One promising radiographic predictor of seizure outcome in mTLE is Apparent Diffusion Coefficient (ADC). ADC is an MRI sequence that is derived from Diffusion Weight Imaging (DWI), and it represents the mean diffusivity of water molecules in tissues. ADC is a marker of tissue degeneration, demyelination, ischemic cell death, and deterioration of tissue integrity in white and grey matter.^10-12^. High intralesional ADC intensity has been observed in TLE patients,^13^ especially in pathological tissues representing as Mesial Temporal Sclerosis (MTS) ^14-16^ and Focal Cortical Dysplasia (FCD)^17, 18^. Thus, it is conceivable that selectively targeting tissues with abnormally elevated ADC signals in the mesial temporal lobe with LiTT may lead to improved seizure control by selectively targeting the most epileptogenic foci.

Past investigations that have demonstrated ADC’s ability to delineate pathological tissues in mTLE, mainly used voxel-wise evaluation. The conventional voxel-wise approach generally does not have the resolution to identify exact spatial positions of high ADC intensity clusters. Moreover, because ADC can be distributed as a gradient across multiple voxels, this approach cannot produce discrete ‘ablation targets’ with clear spatial boundary and morphometry that can be readily incorporated into existing clinical workflow. A method that can identify spatially distinctive targets within the mesial temporal lobe can help optimize trajectory planning and ablation. In this study, we hypothesized that ADC cluster analysis can identify foci of pathological tissues within the mesial temporal lobes of mTLE patients undergoing LiTT. and that the extent of high intensity ADC clusters is associated with seizure outcome.

## Methods

### Subject Information

Thirty patients of the total 66 patients diagnosed with TLE who underwent LiTT at our institution from October 24, 2014, and September 9, 2019, were retrospectively reviewed (protocol approved IRB #00234662). Approximately half of the patients were excluded due to lack of ADC sequence, failure of manual ablation volume segmentation, and accurate co-registration between image sequences. Patients were determined to have unilateral mesial onset mTLE before qualifying for LiTT through video EEG, stereoencephalography (SEEG), or FDG-PET findings. The operative procedure was performed by a single surgeon (WSA) and involved stereotactic placement of a laser fiber (Visualase®, Medtronic, Minneapolis, MN). Real □ time tissue ablation was monitored via intraoperative 1.5 T MR thermometry.

Demographic and clinical characteristics, including intracranial abnormalities, of subjects were studied. Diagnosis of intracranial abnormalities were made based on radiographical interpretation from trained radiologists. Mesial Temporal Sclerosis (MTS) was detected based on T2 flair hyperintensity findings. Focal Cortical Dysplasia (FCD) was identified from stable lesion size over sequential MR images. The 6-month seizure outcomes were collected and categorized based on ILAE scale as employed in previous studies.^19^ A good seizure outcome was characterized as ILAE Class I (complete seizure freedom), and poor outcome as ILAE Class II – VI (aura or seizure present). Patients who were part of the Stereotactic Laser Ablation for Temporal Lobe Epilepsy trial (Medtronic) were excluded from the study.

### Image Acquisition

Preoperative T1 and Diffusion Weight Imaging (DWI) sequence, and intraoperative post-ablation MPRAGE with gadolinium contrast sequence of each patient were collected. The ADC sequence was captured simultaneously when DWI sequence was captured. The preoperative DWI/ADC sequences were capture using echo-planar imaging in a Siemens MRI scanner. DWI acquisition parameters were variable across subjects as reported: Magnetic Field: 3T, b value: 0, 800, 1000 sec/mm^2^, Repetition Time (TR): 4200 - 11100 msec, Echo Time (TE): 52 - 104 msec, Field of View (FOV): 229 × 229 - 245 × 245 mm, Slice Thickness: 2.5 – 5mm, Flip Angle (FA): 90°). The exact distribution of the acquisition parameter is reported in **Table 3**.

### Volumetric Segmentation

Two mesial temporal lobe (mTL) regions commonly targeted in LiTT - hippocampus and amygdala - were automatically segmented from preoperative T1 sequence using Freesurfer Software Suite (Version 7.1.1, MGH) on the ablation side.^20^ T1 sequences and segmented regions were co-registered to the Z - score normalized ADC sequence using Statistical Parametric Mapping (SPM) 12 suite (UCL, 2020). Similarly, intraoperative MPRAGE sequence was co-registered to the ADC sequence using the same approach using SPM12 suite.

After co-registering the intraoperative sequence to the ADC image, the ablation volumes were manually segmented using ITK-SNAP Segmentation Tool from the intraoperative MPRAGE sequence.^21^ The boundaries of the ablation volume were determined by tracing the outer margins of contrast enhancement as per prior investigations.^22^

### Cluster Extraction

After co-registration of mTL volumes, a custom-developed unsupervised clustering pipeline was implemented to identify spatially distinct, hyperintensive clusters in preoperative ADC sequence. Because all subjects had epileptogenic zones localized to mTL areas prior to LiTT, and most ablation volumes were constrained to hippocampus and amygdala, this pipeline was performed exclusively on mTL structures ipsilateral to surgical side. The pipeline identified discrete regions with elevated ADC signals and demarcated them as individual clusters. The spatial boundaries of each cluster were made by iteratively comparing between intra-cluster ADC and extra-cluster ADC signal intensity. The final boundary of each cluster was determined when the intra-cluster tissues had relatively higher ADCs compared to surrounding tissues. The major advantage of this approach is that the ADC threshold to determine the boundary of clusters was dynamic and not predetermined arbitrarily. The pipeline does not require a prior training set or manual intervention because the pipeline produces cluster morphology based on local ADC distributions near each hyperintensive zones,.

The outputs of the pipeline are proposed cluster numbers, volumes of each cluster generated, cluster voxel coordinates, and individual cluster segmentations in native ADC MR space as shown in **Figure 1A** and **1B**. Clusters of sample subjects with good (ILAE Class I) and poor outcome (ILAE Class II-VI) are visualized in **Figure 1C** and **1D**.

**Figure 1.**
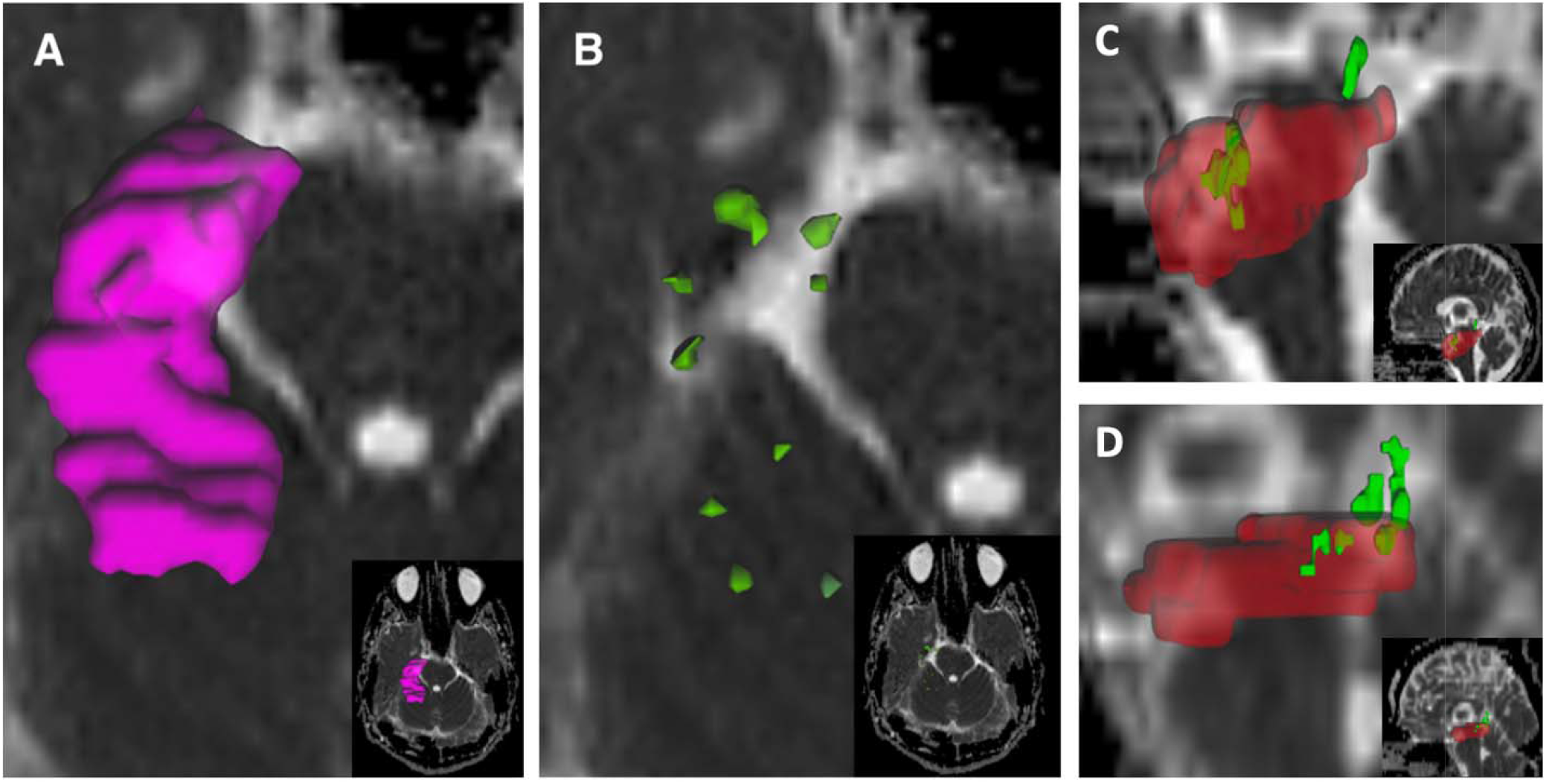
Cluster Extraction and Visualization. **(A)** Visualization of hippocampus (purple**), (B)** The clustering algorithm detects individual, spatially distinct clusters (green) composing the hippocampus. Ablation volume (red) is overlayed onto clusters (green) of sample subject with (**C)** good outcome (ILAE Class I) and (**D)** poor outcome (ILAE Class II – VI).

### Statistical Analysis

#### a. Volumetrics Analysis

For volumetrics analysis, the (1) preoperative whole mTL volumes (cm^3^) and (2) proportion ablated were statistically compared between two outcomes using a nonparametric Mann-Whitney U test. Similarly, preoperative cluster volumes (mm^3^) and the proportion of cluster volumes ablated were compared.

#### b. ADC Analysis

The mean Z-score normalized ADC values of mTL regions were evaluated in preoperative, intraoperative, and postoperative stages between the two outcome categories. This approach was taken to isolate the association ADC may have with the seizure outcome that can be driven either preoperatively based on differential tissue properties, or intraoperatively based on varying ablation regions. The ADC of (1) preoperative whole mTL regions, (2) ablated portion of mTL regions, and (3) postoperative residual mTL regions were compared between the outcome groups. Identical methods were implemented for ADC values in mTL clusters.

ADC voxel intensities have been suggested to be influenced by DWI MR acquisition parameters, such as b-value,^23, 24^ TE,^23, 25^ and TR.^25^ To control for the potential cofounding effect of varying imaging acquisition parameters on ADC intensities, multivariate logistic regression between ADC values and seizure outcomes as binary dummy variables (good outcome = 1, poor outcome = 0) were conducted with DWI MR parameters (b-value, TE, and TR) as covariates. All outliers of ADC values were removed in each outcome group based on median absolute deviation method.

#### c. Cluster Analysis

The validity of the identifying clusters of hyperintense ADC values, as well as targeting these clusters to enhance seizure outcomes, was assessed. First, the number of clusters identified in each of the preoperative mTL regions were compared between outcomes. Second, the proportion of clusters ablated and their association with seizure outcomes were explored.

We attempted to investigate the reliability of cluster ablation by studying the proportion of cluster ablated in two separate ways because generated clusters had relatively small volumes compared to the entire mTL region,: by (1) number and (2) volume. The proportion of cluster *number* ablated was evaluated as the ratio of the number of clusters that have been fully ablated to the total number of clusters proposed. The proportion of cluster *volume* ablated was evaluated as the ratio of the net ablated mTL volumes comprised of proposed clusters to the net volumes of all clusters generated. Employing these different metrics, the proportion of cluster ablated was statistically compared between the two outcome groups.

All statistical tests were conducted using Prism 9 Software Suite (Graphpad Software, San Diego, CA) and MATLAB 2020a (Mathworks, Natick, MA).

## Results

### 1. Demographics and Clinical Characteristics

The clinical and demographic characteristics of subjects are shown in **Table 1**. The mean age at surgery was 36.6 years. Approximately 36.7 % of the subjects had good seizure outcome (ILAE Class I) and 63.4 % poor outcome (ILAE Class II – VI) at last follow-up. Eighty percent of the subjects were diagnosed with MTS, and 6.7% with FCD. Clinical variables such as age at surgery (p = 0.53, Fisher’s Exact Test), sex (p = 0.13, Mann-Whitney U test), handedness (p = 0.13, Fisher’s Exact Test), and ablation side (p > 0.99, Fisher’s Exact Test) were not associated with seizure outcomes. Some subjects have undergone preoperative means to localize the seizure onset zones in mesial temporal structures through SEEG and PET imaging. However, undergoing SEEG (p = 0.37, Fischer’s Exact Test) or having ipsilateral PET hypometabolism findings (p = 0.54, Fischer’s Exact Test) had no association with the degree of postoperative seizure freedom.

**Table 1.**
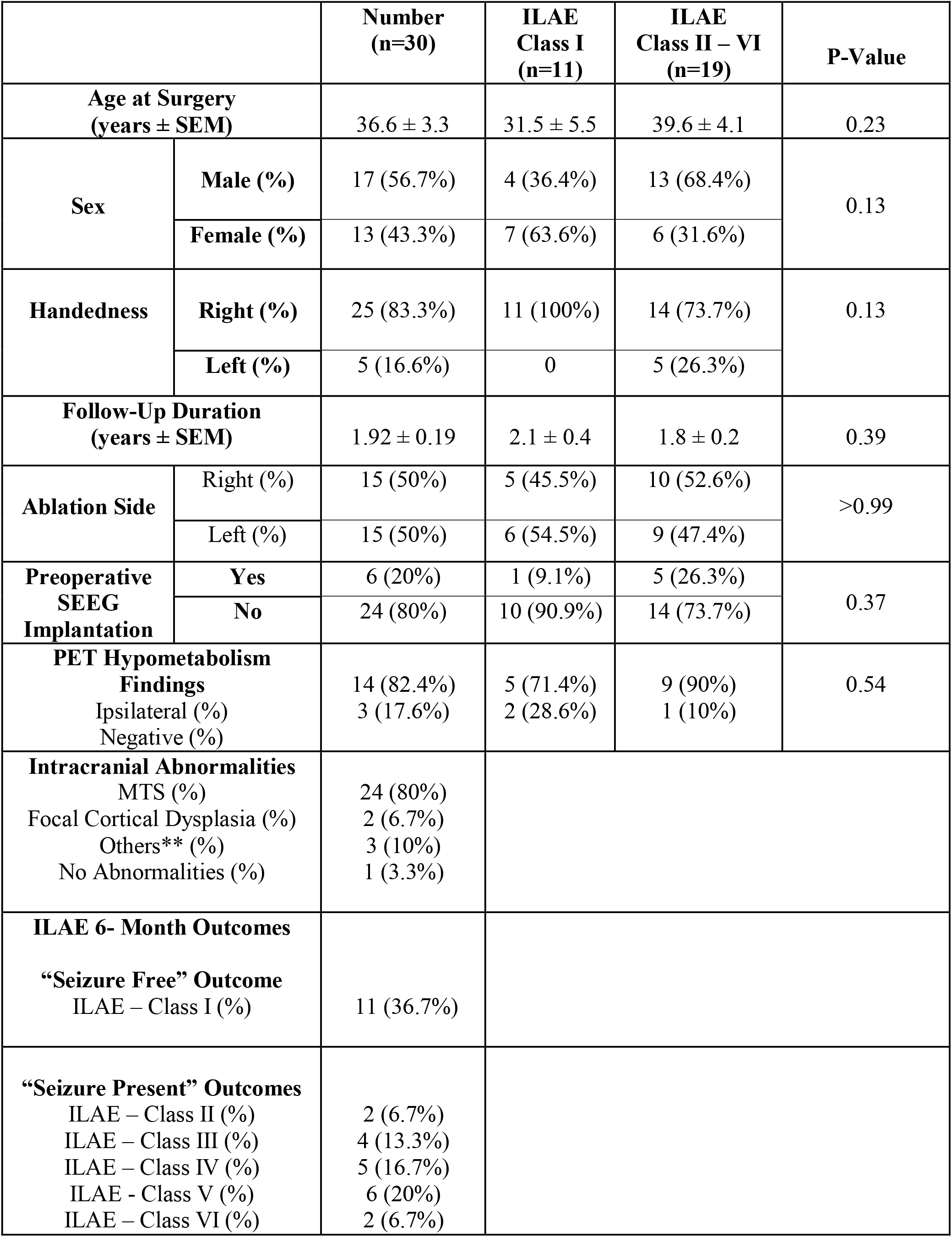
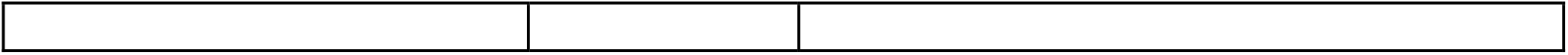
Demographic and Clinical Characteristics. *MTS=Mesial Temporal Sclerosis **Others =Other intracranial abnormalities as hypertrophy or arteriovenous malformation (AVM)

### 2. DWI MR Acquisition Parameters

To account for the variability in DWI MR acquisition parameters used to capture DWI images across patients, statistical tests were conducted to determine the association with seizure outcomes. Three known DWI MR acquisition parameters - (1) b-value, (2) TR, and (3) TE - used to capture ADC map between outcome groups were compared. No statistically significant associations of MR parameters were found.

### 3. Volumetrics

Total preoperative volumes (cm^3^) of hippocampus and amygdala, and each of their proportion ablated (%) was compared between the two seizure outcome groups. Neither the raw volumes of preoperative hippocampus (p = 0.28) and amygdala (p = 0.10), nor the proportion hippocampus (p = 0.12) and amygdala (p = 0.56) were associated with seizure outcomes. Overview of volumetric analysis results is reported in Table 2.

**Table 2:** Whole Structure Volumetrics and Seizure Outcome. The volumetrics values are reported as mean volume (cm^3^) ± SEM.

|  | ILAE Class I | ILAE Class II – VI | P-value |
| --- | --- | --- | --- |
| **Amygdala** |  |  |  |
| Pre-op Volume ($\text{cm}^3$) | $1.79 \pm 0.07$ | $1.94 \pm 0.11$ | 0.10 |
| Total Volume Ablated (%) | $39.83 \pm 8.48$ | $34.27 \pm 4.02$ | 0.56 |
| **Hippocampus** |  |  |  |
| Pre-op Volume ($\text{cm}^3$) | $4.10 \pm 0.25$ | $4.43 \pm 0.22$ | 0.28 |
| Total Volume Ablated (%) | $44.50 \pm 5.92$ | $35.62 \pm 4.06$ | 0.12 |

ADC hyperintense clusters were then extracted using clustering algorithm and pipeline described in **Method 4. Cluster Extraction**. The net cluster volume was defined as a summation of volumes of all clusters found in each amygdala and hippocampus. Cluster ablated (%) was defined as a percentage ratio of volumes ablated comprising the clusters to the net cluster volume. The net volume that summates volumes of all identified clusters within the preoperative hippocampus and amygdala was calculated. The proportion of the net cluster volume ablated in both hippocampus and amygdala was subsequently evaluated. The proportion of hippocampal cluster volumes ablated was significantly associated with seizure outcomes (Mann - Whitney U= 48.50, p = 0.0014, Table 3).

**Table 3:** Cluster Structure Volumetrics and Seizure Outcome.

|  | **ILAE Class I** | **ILAE Class II – VI** | **P-value** |
| --- | --- | --- | --- |
| **Amygdala** |  |  |  |
| Pre-op Net Cluster Volume (mm<sup>3</sup>) | 49.49 ± 15.81 | 59.61 ± 10.11 | 0.25 |
| Net Cluster Volume Ablated (%) | 31.30 ± 7.96 | 19.57 ± 4.70 | 0.21 |
| **Hippocampus** |  |  |  |
| Pre-op Net Cluster Volume (mm<sup>3</sup>) | 82.11 ± 19.38 | 115.58 ± 20.73 | 0.33 |
| Post-Op Net Cluster Ablated (%) | 35.20 ± 5.67 | 16.45 ± 3.69 | **0.014\*** |

### 4. ADC Analysis

#### Whole Mesial Temporal Structures

In the hippocampus, no significant association was reported for ADC of preoperative and postoperative residual hippocampal regions with seizure outcomes. For the ablated portion of the hippocampus, however, subjects with good seizure outcomes had significantly higher ADC than those with poor outcomes (0.13 vs. -0.25, Mann Whitney U = 28, p = 0.008), and the significance held after controlling for MR acquisition covariates (p = 0.016).

In the amygdala, a similar trend was observed and no significant ADC distribution differences between seizure outcomes and preoperative/residual amygdala regions. However, the mean ADC of the ablated portion of the amygdala was significantly higher for subjects with good outcomes (−0.11 vs. -0.48, Mann Whitney U = 9, p = 0.0016), and the significance persisted after controlling for the covariates (p = 0.02).

#### Mesial Temporal Structure Clusters

An identical analysis was performed for extracted clusters from hippocampus and amygdala. For all three cases, no significant association was found between cluster ADC values and seizure outcomes in both univariate and multivariate analysis.

### 5. Cluster Properties and Prediction

#### Cluster Properties

The association between cluster properties and seizure outcomes was characterized by (1) preoperative numbers of proposed clusters and (2) proportion of cluster ablated.

The mean number of preoperative hippocampal clusters across all subjects was 6.2 ± 0.8 (SEM). For subjects with good outcomes, the mean number of proposed hippocampal clusters was 8.3 ± 1.7, and 4.8 ± 0.7 for subjects with poor outcomes as shown in **Figure 2A**. The difference in preoperative hippocampal clusters was not significant (Mann-Whitney U=62, p = 0.067). For amygdala clusters, the mean number of clusters was 2.7 ± 0.3 across all subjects, 3.5 ± 0.7 for good outcome subjects, and 2.3 ± 0.4 for poor outcome subjects as shown in **Figure 2B**. The differences in number were not significant (Mann-Whitney U=67.5, p = 0.090).

**Figure 2.**
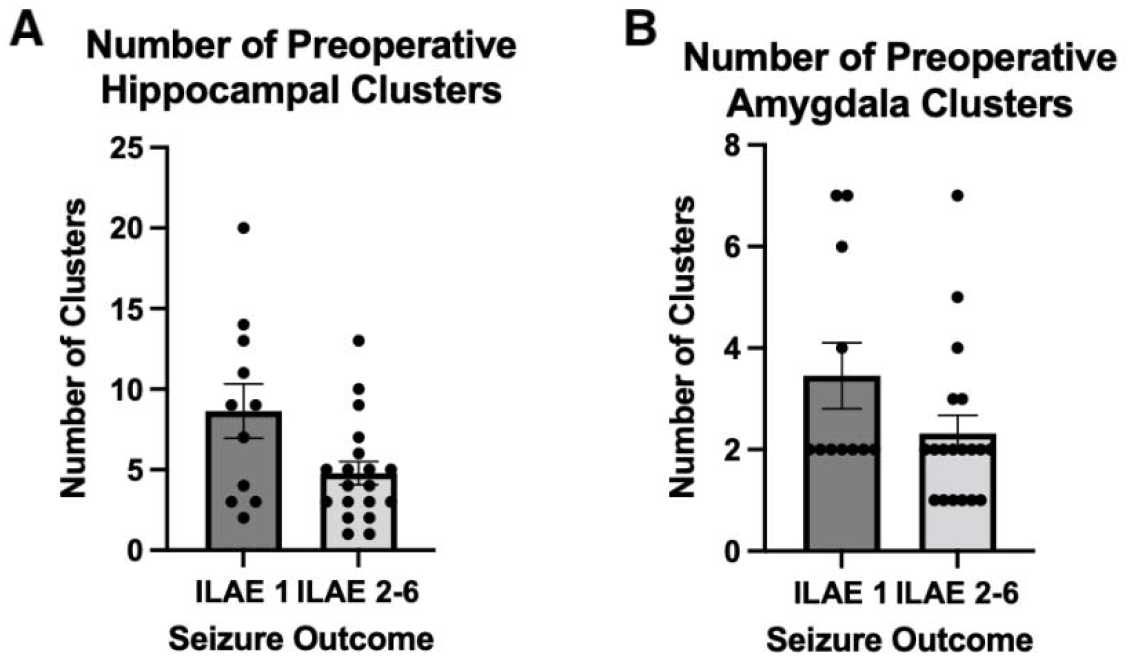
Preoperative Cluster Numbers and Seizure Outcome. The association of seizure outcome is between hippocampal **(A)** and amygdala **(B)** clusters.

For hippocampal ablation, the mean proportion of hippocampal cluster number ablated across all subjects was 12.6 ± 3.5 %. Subjects with the good outcome had higher proportion ablated than subjects with poor outcome (27.1 % vs, 4.2 %, Mann-Whitney U=40, p = 0.0007). For amygdala ablation, the mean proportion ablated across all subjects was 5.2 ± 2.7 %, and subjects with the good outcome also had a significantly higher proportion of amygdala ablated (13.2 % vs. 0 %, Mann-Whitney U=59.5, p = 0.016). Results of proportion of hippocampal and amygdala cluster numbers ablated are reported in **Figure 3**.

**Figure 3.**
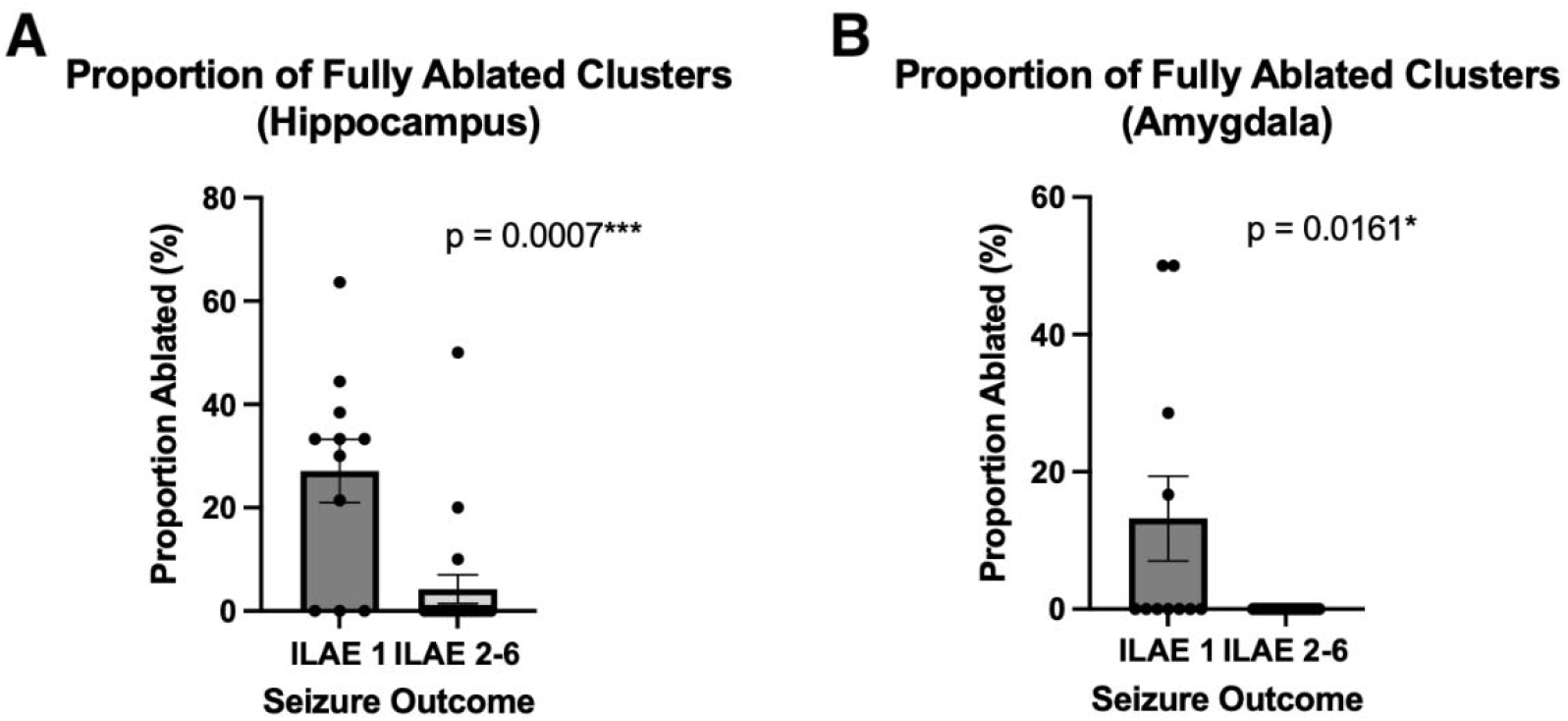
Proportion of Fully Ablated Clusters and Seizure Outcome. Mean proportion of cluster numbers ablated for hippocampus **(A)** and amygdala **(B)**.

#### Hippocampal Cluster Ablation and Seizure Freedom Prediction

The predictive power of hippocampal cluster ablation on seizure outcome was assessed. Using univariable logistic regression, the proportion of cluster ablated was fitted with binary seizure outcomes (ILAE Class I = 1, ILAE Class II – VI = 0). The predictive power of the cluster ablated was independently analyzed based on two different metrics: (1) by net cluster volume and (2) by cluster numbers ablated.

Based on univariate logistic regression between proportion of cluster ablated and seizure outcome, seizure outcomes was significantly associated with proportion of cluster ablated based on cluster volume (OR = 1.063, 95% CI [1.016 – 1.126], p = 0.017), and by cluster number (OR = 1.084, 95% CI [1.031 – 1.616], p = 0.0063). Based on the AUC of the ROC curve, the predictive capacity of both of cluster ablation metrics also proved to be significant. The AUC of ROC between cluster volume ablated and seizure outcome was 0.7679 (95% CI [0.58 – 0.96], p = 0.0160). AUC of ROC for cluster number ablated was 0.8086 (95% CI [0.6258 – 0.9914], p = 0.0055). ROC curves based on proportion of cluster ablated by volume and by number are shown in **Figure 4A** and **4B**, respectively.

**Figure 4.**
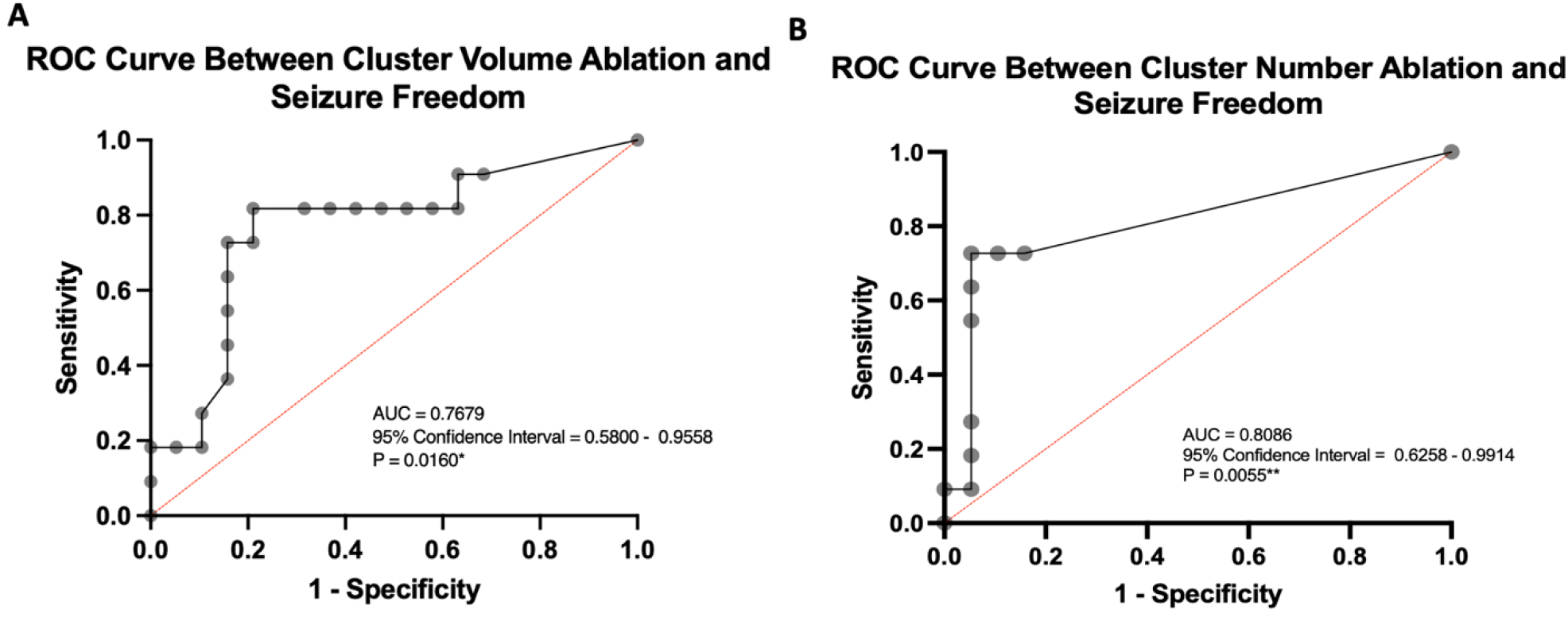
ROC Analysis for Seizure Freedom from Hippocampal Cluster Ablation. ROC curves were generated from univariable logistic regression between seizure outcome and degree of (A) cluster volume and (B) cluster number ablated.

## Discussion

LiTT is a highly promising surgical alternative to anterior temporal lobectomy. However, reported seizure outcomes after LiTT are highly variable despite efforts to standardize the procedure. One possible reason is that the current LiTT techniques do not adequately address patient variability; Hence, there is a need for identification and validation of novel patient specific factors that are predictive of seizure outcome. ADC is a radiographic marker of disease that is easy to acquire and incorporate into the clinical workflow, can enable surgeons to fine tune the laser trajectories and intra-operative post-ablation assessment to ultimately enhance seizure outcome.

Our study explores the utility of (1) identifying hyperintense ADC clusters within mesial temporal structures and (2) ablation of these clusters improves mTLE seizure outcome following MRg-LiTT. Employing a machine-learning based approach, spatially distinct, discrete clusters were identified in the hippocampus and amygdala. The degree of hippocampal clusters ablated, both by volume and number, was significantly associated with complete seizure freedom (ILAE Class I) and was able to reliably predict seizure outcome following LiTT. To the best of our knowledge, this is one of the first studies to evaluate the relationship between ADC clusters in the mesial temporal structures and seizure outcome in mTLE patients following LiTT.

In mTLE, pathology is generally localized in the mesial temporal structures, and primarily in the amygdala, hippocampus, subiculum, and parahippocampal complex, that are strongly associated with seizure generation and initiation.^26-31^ We chose ADC as a surrogate marker of pathological tissues distributed across the mesial temporal structures. ADC measures have been closely connected to mTLE pathology, as well as in comorbid tissue abnormalities as MTS and FCD. Elevated ADCs have been reported in ipsilateral mesial temporal structures in the ictal side,^32, 33^ and tissues composed of MTS,^34^ and FCD.^35^ While it is unclear whether the tissue clusters consisting of distinct ADC hyperintensity represent the highly epileptogenic zones, mounting evidence suggests that formation of pathological tissues observed in MTS and FCD may be a consequence of chronic seizure generation.^36-39^. These local pathological tissues may be detected based on radiographic evaluation, such as in MTS ^40, 41^ and FCD,^42, 43^ or based on abnormal electrophysiological signals detected with EEG or SEEG.^44^

In a previous study, we demonstrated that mTLE patients with higher ADC values within the ablation zone had significantly higher chance of good seizure outcome.^45^ In this study, our results further suggest that the clusters likely demarcate patient specific pathology that are epileptogenic and/or is part of the main seizure network. Also, the fact that we failed to find association between proportion of hippocampus or amygdala ablated and seizure outcome also suggests that currently used ablation volumetrics may be inadequate because it fails to account for the heterogeneity of epileptogenic zone volume and locations among patients.^46^

According to our findings, inter-cluster ADC values of the amygdala and hippocampus were not associated with seizure outcomes. Furthermore, no difference in the proposed number and volumes of clusters was observed between the seizure outcomes. These collectively support the consistency of the clustering approach to producing distinctive clusters. Through this approach, the proposed numbers, size, and ADC intensities forming the clusters have been controlled for, and seizure outcome became dependent only on the proportion of clusters ablated.

For the case of volumetrics, the total amygdala and hippocampal cluster volumes proposed were approximately 54.5 mm^3^ and 98.8 mm^3^, respectively. In meta-analysis across 13 studies, the pooled mean total ablation volume was 5376 mm^3 47^ with a range of 2900 – 7110 mm ^3^.^48, 49^ Relatively smaller cluster volumes allow for more focused ablation targets, which may allow for a reduction in potential postoperative cognitive complications. Compared to traditional open resection procedures, it is reported that cognitive measures, such as object naming and recognition,^5^ are preserved after laser ablation. It is presumed that a more favorable outcome is expected due to more focal tissue damage without affecting other regions involved in cognitive processing,^50^ including extra-mesial temporal regions. Additionally, a decrease in verbal memory and visual memory was observed with the extent of left and right temporal lobe resection, respectively,^51^ in temporal lobectomy. Through our approach, it is may thus conceivable to further preserve cognitive functions with more precise means for targeting.

From our results, the proportion of (1) cluster volumes and (2) cluster numbers ablated were significantly associated with seizure outcome. For the hippocampus, the increasing degree of both cluster volumes and numbers was correlated with complete seizure freedom. For the amygdala, only a reduction of cluster numbers was associated with seizure freedom. Acknowledging the topographic variability between the amygdala and hippocampus, aiming to ablate the maximum number of spatially distinct clusters detected may pose a substantial therapeutic effect. In the case of hippocampal clusters, we further established a strong predictive power of cluster ablation on seizure outcomes through ROC analysis on both number (AUC = 0.8086) and volume (AUC = 0.7679).

With an increasing understanding and growing works of literature on potential predictors of LiTT outcome, we here propose that significant seizure freedom may be expected when clusters of hyperintense ADC signals are targeted. This information may be crucial for presurgical planning, such that trajectory and laser power can be adjusted to produce ablation volume maximally composed of pre-operatively detected clusters. Further benefits may also extend to the postoperative reduction of neuropsychological complications due to relatively small, proposed ablation volumes.

## Limitations

This study has several limitations. First, it is a retrospective study with a relatively small cohort size. Our 6-month postoperative seizure freedom rate was 36.7%, which was lower than that previously reported rates, which suggest difference in patient population or selection criteria for LiTT among centers.^52^ Prospective multicenter investigation with a larger population and longer follow-up is needed to validate our results.

Second, variability in acquisition parameters of the DWI / ADC map as all images were extracted from a retrospective clinical database. Because voxel intensities of ADC sequence are known to be sensitive to parameters such as b-value, TR, and TE, we acknowledge the potential confounding effects when analyzing ADC signals. We attempted to minimize these effects and extend the generalizability of our findings by undergoing a pre-processing step and normalizing the voxel intensities (Z-score) of each ADC map. Standardization of the voxel intensities for each sequence partly allowed us to compare them across the two clinical cohorts and use them to produce more reliable clusters. Furthermore, we further attempted to control for acquisition parameters by including them as beta coefficients in multivariate analysis when comparing ADC signals between cohorts.

Third, we performed ablation volume segmented based on intraoperative post-ablation sequences. Because past studies have suggested that ablation volumes tend to decrease following initial ablation,^53^ it is possible that our approach may have overestimated the ablation volumes. However, we have had restricted access to delayed post-operative MRI because our institutional clinical protocol only captures intra-operative T1 MRI sequence. Furthermore, only a few subsets of subjects had undergone post-operative MRI because it is generally performed for those with post-operative recurrent seizures. Therefore, we used intraoperative sequence for this study because it was the only sequence available in every patient in our cohort. The use of intra-operative post-ablation sequence to segment ablation volumes is also consistent with methods employed in previous literature on large multicenter study.^52^ Future study should evaluate mTL volumetry with ablation volumes using delayed post-operative imaging.

## Conclusion

Greater extent of ablation of ADC hyperintensity cluster in the hippocampus, and the amygdala to some degree, was significantly associated with complete seizure freedom in mTLE patients who underwent LiTT. Our results suggest that ADC may potentially identify patient-specific pathological and epileptogenic areas in the mesial temporal lobe that are high yield for ablation. ADC cluster-based approach can enable clinicians to optimize patient selection process, pre-operative planning, and intra-operative post-ablation assessment to further improve seizure outcomes.

## Data Availability

No data produced in the present work are publicly available.

## Acknowledgements

None

